# *“They provide the right kind of support.”* A qualitative study of preferences for differentiated service delivery location among recipients of antiretroviral therapy at Lighthouse Trust in Lilongwe Malawi

**DOI:** 10.1101/2023.12.16.23300076

**Authors:** Lisa Orii, Kate S. Wilson, Jacqueline Huwa, Christine Kiruthu-Kamamia, Odala Sande, Agness Thawani, Astrid Berner-Rodoreda, Evelyn Viola, Hannock Tweya, Petros Tembo, Wapu Masambuka, Richard Anderson, Caryl Feldacker

**Affiliations:** Paul G. Allen School of Computer Science & Engineering, University of Washington, Seattle, Washington, United States of America; Department of Global Health, University of Washington, Seattle, Washington, United States of America; Lighthouse Trust, Lilongwe, Malawi; Faculty of Medicine, Heidelberg Institute of Global Health, Heidelberg University, Heidelberg, Germany; International Training and Education Center for Health (I-TECH), University of Washington, Seattle, Washington, United States of America

## Abstract

Differentiated service delivery (DSD) models for antiretroviral therapy (ART) allow stable recipients of care (RoC) to receive multi-month ART drug refills and complete rapid reviews in community sites. As DSD options expand across sub-Saharan Africa, RoC’s preferences and perspectives on community-versus clinic-based care models warrants attention. We describe the factors that influence RoC choice of ART delivery approaches at Lighthouse Trust (LT) clinics and community-based DSD sites in Lilongwe, Malawi. We conducted eight focus group discussions (FGDs) among LT RoC in the Nurse-led Community-based ART Program (NCAP) (n=4) and in clinic settings (n=4) to explore opinions, preferences, and perceptions about ART service delivery. FGDs were conducted and recorded in Chichewa and then translated and transcribed into English for analysis. Data was analyzed using thematic analysis and findings discussed with the LT and NCAP teams to jointly reflect on the findings. Sixty-three participants took part in the qualitative study. Results were largely similar across care locations. In both NCAP and clinic FGDs, RoC appreciated the convenience of integrating their appointment visits at their chosen care location into their daily lives, though some RoC traveled far to access LT’s high quality of care. RoC were satisfied with the quality of the care they received at their location of choice. Privacy protection was an important consideration for choosing care models. At LT clinics, RoC highlighted the importance of physical separation between LT’s HIV-specific service site and other service sites. In NCAP, RoC expressed that their choice of care model was reinforced by the sense of mutual support that they received through the peer support model. At LT, RoC in both clinic and NCAP care models expressed satisfaction with their chosen care model and preferred that choice over alternative options and locations. Overall, LT RoC appreciated the quality of care, the respectful provider-to-patient interactions, and the attention to privacy at community and clinic sites. These findings suggest continued emphasis on offering choices to RoC on where and how they receive ART delivery approaches may support ongoing engagement in care.

## Introduction

To reach the global target of eliminating new infections by 2030 [1], health systems in Sub-Saharan Africa (SSA) must adapt existing resources to increasing numbers of recipients of care (RoC) on lifelong HIV care and treatment. Malawi has 950,000 adults aged 15 and over living with HIV (7.1% prevalence), of whom 92% receive antiretroviral therapy (ART) [14]. Since 2017, many countries have adopted differentiated service delivery (DSD) models, where RoC with suppressed viral load and good antiretroviral therapy (ART) adherence can receive multi-month drug refills to reduce clinic visits and receive ART services in community sites [2]. DSD models were designed to be more client-centered and efficient than standard monthly clinic visit models, although there is limited evidence on comparative feasibility, quality, or effectiveness between different models or settings [2,3]. One randomized trial in Zimbabwe found that retention and viral suppression outcomes were similar between RoC receiving 3-month refills from community sites compared to those receiving 3-month refills at clinics, suggesting that community-based ART models are as effective as clinic based extended refills [4]. Recent qualitative studies explored RoC and provider experiences in HIV care with DSDs. RoC in community-based models reported that fewer clinic visits reduce their clinic wait times and transportation costs [5–7], increase peer support [8,9] and improve treatment adherence [10]. By contrast, clinic-based RoC in SSA reported preferring clinic-based care to community-based refill options because the clinics were near work [5] or they feared stigma and discrimination of inadvertent disclosure of their HIV status if they collected ART at a community venue [5,6,8,11,12]. Healthcare providers also reported that RoC’s concerns around confidentiality and unwanted disclosure of HIV status posed barriers to using clinic-based DSD models [5]. It is possible that RoC may have different reasons for accessing ART care at clinics or community sites, and perspectives from RoC in routine HIV care with both clinic and community-based DSD options are lacking.

Given national efforts to expand DSD options in SSA to improve RoC-centered care in clinic and community sites [13], RoC preferences, opinions, and perspectives on care models warrant further attention. To address this gap, we conducted a qualitative study among RoC to explore opinions, preferences, and perceptions about ART service delivery locations among both community- and clinic-based RoC at Lighthouse Trust (LT) in Lilongwe, Malawi, a large, public ART provider with over 38,000 RoC in two urban clinics and 120 community-based care sites. This qualitative study may inform service delivery considerations in other low-resource ART programs in the region.

## Methods

### Setting

LT supports the ART program of the Malawi Ministry of Health and provides integrated HIV prevention, treatment, care, and support services in five high-volume Center of Excellence clinics across the country. LT is widely recognized for its innovations and efforts to improve the quality of care over two decades of HIV-related service delivery [15–22]. In urban Lilongwe, LT runs two, large flagship clinics with 25,148 RoC at Martin Preuss Center at Bwaila Hospital and 12,853 at the Lighthouse clinic at Kamuzu Central Hospital. At these flagship clinics, viral load suppression at 12 month’s ART averaged >90% across all sites according to routine monitoring and evaluation data,. LT also supports care at 9 peri-urban satellite sites in Lilongwe district. LT implements the same training, policy, and practices at all sites, including use of the Malawi Ministry of Health’s real-time, point-of-care electronic medical record system. LT implements several DSD approaches in its clinics, including expanded early and late service hours, adolescent-friendly programs, integrated family planning services, and non-communicable disease care. LT also operates a large-scale, community-based DSD model, the Nurse-led Community-based ART Program (NCAP). In NCAP, Community Nurses provide ART services to eligible RoC referred from all clinics. RoC from LT clinics may be referred to NCAP if they meet the following criteria: 1) at least 18 years of age; 2) clinically stable; 3) on ART for at least 6 months; and 4) member of a peer support group. All NCAP clients previously received services at LT clinics. NCAP is provided in the community during routinely scheduled peer support meetings held in churches, community centers, schools, or other gathering places according to the peer group preference. RoC meet with a community nurse at that same location every 3-6 months. In 2023, over 5,195 RoC in 120 groups in Lilongwe were enrolled in NCAP and reported viral load suppression among NCAP RoC on treatment was ∼95%. NCAP receives high levels of client satisfaction and healthcare worker support for expansion [6].

### Study design and population

As part of a larger qualitative study on data privacy and security in both community and clinic settings [23], we conducted eight focus group discussions (FGDs) with LT RoC at both flagship and NCAP sites from September 7, 2022 to October 14, 2022. The facilitator was a trained Malawian LT qualitative researcher who speaks both Chichewa (the most common language in Malawi) and English. We used purposeful sampling through clinic and community-based care locations to 4 FGDs per setting. LT staff in both settings screened RoC study eligibility: 1) age 18 and above; 2) at least a primary education; 3) written voluntary consent to FGD participation; and 4) permission to audio record the discussion. FGDs were conducted at participants’ routine care location, either the clinic or NCAP site.

### Data collection

Four FGDs took place at clinics and four in NCAP sites. Each FGD included seven to nine participants. FGDs were conducted using a semi-structured discussion guide. The guide explored RoC’s experiences with health care in their preferred care location. In this study, we focus on reasons for accessing ART services at their chosen care model and consideration for alternative care locations. A Malawian qualitative researcher facilitated all FGDs in Chichewa. FGDs were audio recorded with consent from all participants and lasted about an hour. After each FGD, the facilitator translated and transcribed the FGDs into English and completed a debrief report summarizing key points, participants’ openness, and group dynamics. The research team reviewed and discussed all debrief reports.

### Analysis

FGD transcripts were analyzed using thematic analysis, combining deductive and inductive steps [24]. Two team members (located in the United States and who did not conduct the FGDs) conducted a detailed reading of the full-length transcripts and developed an initial codebook restricted to the questions about care model preferences, including concepts related to stigma, in ATLAS.ti v.8. A second team member reviewed the code application and resolved disagreements through discussion with the main analyst. Final coding, discussion, and next steps were refined through highly participatory discussion between the analysis team and LT study members.

### Ethical considerations

This study was approved by the University of Washington Institutional Review Board (STUDY00013936) and the National Health Sciences Research Committee of Malawi (# 2968). All participants provided written informed consent. All participants were compensated with K10310.00 (approximately 10 USD) for their time. This study was conducted in accordance with the LT team and researchers whose primarily interest is to improve the quality of care of RoC.

### Positionality

The author group consists of international and local researchers. The group benefited from in-depth local knowledge and experience of HIV studies across various geographic settings. All authors were attentive to iterative process of data review and reflection [25] to strengthen the study’s methodological rigor, transparency, and inclusion throughout the research process, especially during data collection, analysis, and results reporting.

## Results

Overall, 63 participants took part in this study. The median age was 47 (IQR 41-56), 81% were female, 51% received care from NCAP and 49% from clinics (Table 1). Overall, participants from both NCAP and the clinics reported positive experiences with their chosen care model. We identified four themes about participants’ reasons for choosing their care model: (1) convenience including integration into daily activities and efficiency; (2) high quality of care; (3) assurance of privacy; and (4) sense of mutual support. We compare findings from clinic and NCAP discussions.

**Table 1.**
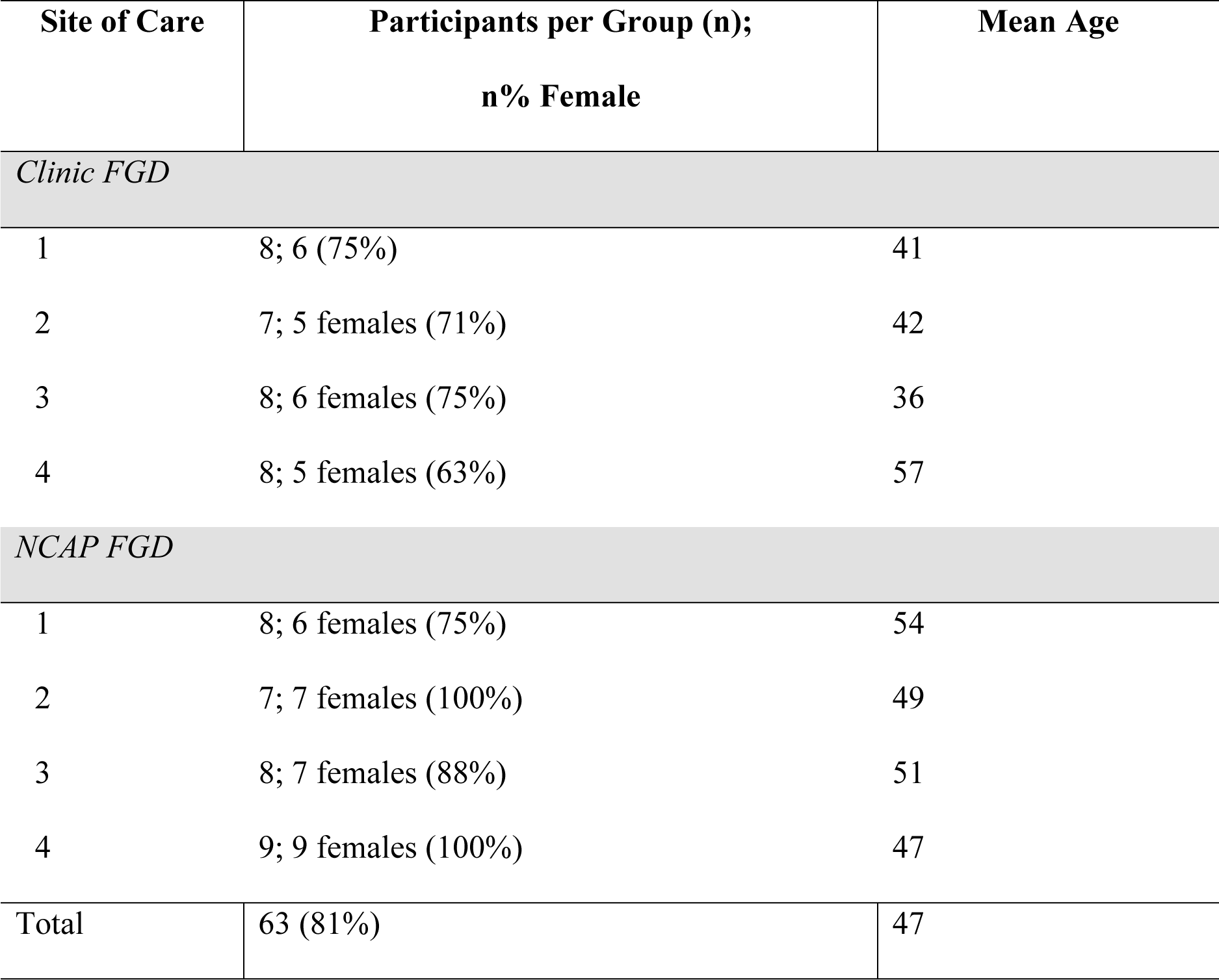
Gender and Age Distribution of FGD Participants.

| Site of Care | Participants per Group (n);<br>n% Female | Mean Age |
| --- | --- | --- |
| *Clinic FGD* |  |  |
| 1 | 8; 6 (75%) | 41 |
| 2 | 7; 5 females (71%) | 42 |
| 3 | 8; 6 females (75%) | 36 |
| 4 | 8; 5 females (63%) | 57 |
| *NCAP FGD* |  |  |
| 1 | 8; 6 females (75%) | 54 |
| 2 | 7; 7 females (100%) | 49 |
| 3 | 8; 7 females (88%) | 51 |
| 4 | 9; 9 females (100%) | 47 |
| Total | 63 (81%) | 47 |

### Recipients of care wanted convenience

The convenience of integrating appointment visits into daily routines influenced participants’ choice of care model. This sentiment was expressed more strongly among participants from NCAP than the clinics. Short travel distance was a major factor that influenced participants’ choices.

> *“I am thankful for this arrangement where we must be getting medication in the community. Sometimes we leave some activities in the community, we come here and get medication and attend to activities like funerals within the community. In addition, when we are sick, we can get medication here when the providers visit the community…Bringing the community ART program has helped us a lot. We can leave beans boiling on fire, come here and get back home, and still find the beans boiling. We are very grateful.”* – P5- NCAP2

For some clinic-based participants, short distance to the urban clinic and extended hours of operation enabled them to integrate clinic visits into their work schedule.

> *“I like this clinic because it is near to my home, as such I do not struggle with transport. In addition to that, they open the clinic early in the morning. I can go back to work once I collect my medication.”* – P1-Clinic2

Many NCAP participants compared their current travel to their previous clinic-based care, saying that the distance, travel time, wait time, and costs associated with accessing services reduced after opting into NCAP. Further, participants reported that lower costs helped them attend support group meetings and receive consistent ART.

> *“Another thing is about transport, some fail to report to the clinic because they do not have means of transport and cannot walk to the clinic, they end up missing appointment dates there by not being able to get medication. Getting medication in the community has helped us a lot because we get here easily, we can just walk to the site.”* – P6-NCAP2

### Recipients of care valued high quality of care

In FGDs from both clinics and NCAP, RoCs expressed that providers respected and cared for them kindly, which affirmed their choice of care location. At the clinics, participants noted that nurses offered adherence counseling and emotional support to encourage them to live healthy lives and accept their serostatus.

> *“Since I started taking medication. I am encouraged especially because of the counseling that is provided at Lighthouse so that one accepts their status. They provide counseling on being adherent and other issues that make one encouraged and be healthy (as it was before).”* – P1-Clinic3

Clinic-based participants also compared the quality of care at LT clinics to that of other hospitals or clinics, making informed choices on which clinic to attend:

> *“There are some sites where one would be disappointed because they have been shouted at. Even though I live far from here, I make sure that I report to the [Lighthouse] clinic”* (P3-Clinic2).

Distance was therefore not a major consideration for some RoC when choosing care locations.

> *“Even though I live far from here, I heard that they manage the disease very well at this clinic. When I was referred here, I noted that there is expertise at this clinic. As I continued collecting medication here, I noticed that they have good reception. During my first visit, I noted that there is respect and love for the client, this is different from what I experienced in other sites. It made me happy, and I decided that even though I live far from the clinic I will be reporting to this clinic.”* – P6-Clinic2

NCAP participants reported that community nurses were more respectful and friendly as compared to clinic providers. This experience reflected a common perception that NCAP providers were interested in the lives of RoC, which encouraged RoC to openly share health issues with nurses. One participant characterized NCAP providers by noting that,

> *“they warmly welcome us; they are happy and interested in us. In addition to that they are attentive when we are sharing our issues with them, they provide the right kind of support”* (P6-NCAP4).

Providers offering responsive and appropriate care that was easy to understand was seen as another incentive for staying in NCAP. Specifically, participants appreciated how NCAP providers offered referrals and reminders for other preventative healthcare.

> *“Our providers provide services appropriately, whenever we have issues in addition to the medication that we get, they act so fast to refer one to the clinic where they can get assistance. Whenever they have something to share with us, they make sure that they deliver the information in a proper manner so that we can understand. We are so happy with the doctors who come here.”* – P4-NCAP2

### Recipients of care wanted their privacy protected

Privacy was an important consideration for all participants in choosing their care model. Clinic-based participants chose longer travel times and distance to receive care at LT to avoid encountering people from their communities. The primary reason was fear of unwanted disclosure of HIV status and discrimination.

> *“I also come from far, but I still come here because I believe my privacy can be maintained because it is far from my home. I did not meet people from my community when I came here. There are others who discriminate against others once they learn their HIV status. When I came here my confidentiality is maintained.”* – P4-Clinic2

Many participants reported that they felt reassured that they would not be seen by people they knew because the LT ART clinics are in a separate section of the hospital.

> *“I am happy, nowadays. Those of us who are positive and on treatment are stigmatized, we are seen differently. I am happy because they separated the clinic, it isolates us from them, and they do not know where we go and what we do there. It makes me happy to come to this clinic.”* – P5-Clinic1

Within the LT clinic, participants felt that confidentiality was well-maintained, especially through the privacy that was given during conversations between RoC and providers.

> *“When we get into the nurse’s room, they tell us to close the door. It is only the client and the nurse in the room. So, whatever we discuss is confidential, it remains between the nurse and the client. They maintain our confidentiality.”* – P2-Clinic3

Conversely, several NCAP participants reported they had more privacy at NCAP sites. One main reason for this was the proximity between a LT clinic and other service areas of the larger hospital campus that offered non-HIV services. Some participants expressed discomfort with being seen visiting LT by others including guardians sent to LT by fellow RoC. Consequently, fear of unwanted disclosure of positive HIV status deterred some RoC from reporting to the clinics for their appointments.

> *“Sometimes it happens that one has sent a guardian, and you meet them (referring to the guardian). It then becomes embarrassing when you meet at home. If the person that you met is not able to keep it to themselves then they will tell other people that you met at the clinic and that you are taking medication. That made others not to report to the clinic with fear that they will meet other people who know them.”* – P8-NCAP3

RoC who chose NCAP also explained that physical separation of spaces at NCAP sites enhanced the sense of privacy.

> *“When the nurses get here, there is a distance between them and the recipients of care, even if one looks at them there is nothing that they can hear. There was another place where we had a side room, where one would not hear what is being discussed no matter how hard they try to pay attention. There is a distance between other recipients of care and the nurse.”* – P3-NCAP4

However, some RoC chose to remain in clinic-based care because of the risk of unwanted disclosure through NCAP participation.

> *“In the past there were groups and once you join you were connected to a smaller group where you were visited. But there were the same people (from the group) who were disclosing other people’s status, yet this is confidential. A lot of people thought that those groups were not good, and a lot of people decided to drop out.”* – P7-Clinic4

### Choice of care model reinforced by sense of mutual support

There was appreciation for peer support exclusively among NCAP participants. The sense of mutual support expressed by NCAP participants was less related to the physical location of the NCAP sites. Most NCAP participants reported that the community group model enabled them and their peers to stay better engaged in care compared to at the clinics. Participants shared how they supported each other to attend community visits, including volunteering to call or visit other RoC to remind them to meet.

> *“We are very comfortable. We also encourage each other as clients who report to the community to get medication. We encourage each other that if we have noticed one of our friends is sick and is not aware of the main issue, we visit them. Though we are not volunteers, we tell them the facility where they can get help. It is good to be open.”* – P4- NCAP2

The mutual support between NCAP RoC also enabled them to encourage one another to seek care for other health issues.

> *“We are happy to be getting medication in the community. It is a good thing because we encourage each other about health and some of the issues that we face. Unlike at the clinic like [X] or [Y] where there are different people, and it is hard to be free to interact. Here [in the community] we meet too often, and it makes it easy to encourage each other.”* – P6- NCAP2

NCAP participants also felt that they supported the nurses’ role in care delivery. NCAP RoC and group leaders worked closely with community nurses to track other RoC who missed their scheduled appointments, which likely helps to reduce the number of defaulters. The collaboration between the group leader and the nurses also allowed volunteers to deliver medication to RoC who, for various reasons could not collect them from the nurse, thus encouraging continuation of care.

> *“We work with the providers, and this has reduced the number of defaulters…We are able to know the appointment dates of the clients and we are able to remind them, when they do not show up to the community distribution point. We follow them up and bring them here. It is unlikely to have defaulters and we have reduced the workload for the providers.”* – P7-NCAP2

## Discussion

In this qualitative study to explore RoC preferences for community- or clinic-based ART care models, we found similar reasons for why both groups chose their community or clinic-based model reinforcing the value of continuing to offer multiple care settings. Convenience, privacy, and quality of care were perceived as facilitators for both groups in selecting their preferred care model. Those in NCAP also commented on the role of peer support in improving their care experience. Choice of location offered RoC with the flexibility to choose the care model that best meet their needs, likely boosting satisfaction that could improve motivation to remain in care. Comparing perspectives of RoC on ART services in two settings run by the same clinical team enhances the strength of the results. These findings suggest several areas of consideration.

RoC in both locations were concerned with privacy and the potential for stigma, making a deliberate choice to remain in their care location to avoid HIV-related stigma. While DSD is scaling up, frequently to community settings, previous studies suggest fear of unintended disclosure as a barrier to uptake of community-based DSD models [5,6,8,11,12,26]. In our study, some clinic-based RoC compromised travel and time concerns to receive care at LT, rather than in the community, to avoid being seen receiving HIV-related services. The separation of LT ART clinics from other sections of the hospital enhanced clinic-based RoC’s sense of privacy. While this may be unique to the operation of LT, physical separation of ART clinics from the main facility likely contributed to the sense of privacy and comfort. These results again reinforce the need for diverse care setting options.

RoC appreciated care models that accommodated their daily routines, underpinning the importance of designing care models that address the needs and preferences of RoC [30]. RoC from both clinics and NCAP recognized the importance of integrating appointment visits into their daily routines. Clinic-based participants appreciated that they could easily fit their visits into their work schedules,. For NCAP RoC, the proximity of NCAP sites to their homes allowed them to participate in community activities and household chores.

Quality of care was another important consideration in participants’ choice of care model. Clinic-based RoC reported high satisfaction with the static-site teams, which may be attributed to the comprehensive and advanced care options provided by the LT clinics, allowing easy access to integrated high-quality care in a single visit. Although some RoC in community-based models may fear detachment from the formal health system [11,31], NCAP RoC felt that NCAP enabled comprehensive care through referrals and reminders for other preventative healthcare made by community nurses. Even more, NCAP RoC preferred the quality of treatment they received from community nurses over that of clinic nurses, noting their friendliness and respect. Community nurses attend to few RoC overall, allowing for a more tailored and personalized care approach. RoC satisfaction with their interactions with community nurses supports the potential for improving RoC-centered care with DSD models [32].

This study complements previous NCAP pilot evaluation [6] that identified the central role of peer support in supporting health and wellness among NCAP participants. NCAP RoC felt that community-based care brought individuals together for peer support, encouraging them to overcome barriers to retention. Peer support groups with strong leaders like in NCAP may also minimize the nurse workload, reflecting potential DSD advantages for both providers and RoC [33, 34]. In this study, participants continued to appreciate reduced costs and improved convenience of NCAP, appear satisfied with their care, and value the privacy benefits afforded by community-based care. However, previous NCAP evaluation noted LT crowding as a primary reason to switch to NCAP-based care, something that was not identified in this study. NCAP expansion may have reduced LT wait times, an intended positive result of DSD scale-up.

This study also had limitations. First, the flexibility that is offered with NCAP participation is restricted to eligible LT RoC, not all LT clients. We engaged participants established in care and their reasons for choosing NCAP or clinic-based care are subject to recall bias. There is potential loss of nuance in participants’ contributions in translating FGDs from Chichewa to English. We tried to minimize this with the involvement of a Malawian qualitative researcher who conducted and translated the FGDs into English. Despite limitations, these findings offer useful feedback for program planners and providers.

## Conclusion

RoC are making informed choices about where to receive care, considering convenience, quality of care, protection of privacy, and potential for mutual support. The varying needs, characteristics, and preferences of RoC underscore that there is no one-size-fits-all care model, but rather that lifelong HIV care requires flexibility for RoC to choose between clinic-, community- or other DSD-type models as preferences and life circumstances change. This adaptive and responsive approach can reduce burdens and concerns that impede access to services and retention in care. Although not all care options are always available to everyone across locations, increased flexibility for care model changes and allowance for more fluid movement between models may better meet RoC expectations for convenience, privacy, and quality care, enhancing retention and satisfaction over time.

## Data Availability

Data cannot be shared publicly because of privacy concerns among people living with HIV at this clinic who may be identified by findings of the focus group discussions. Data are available from the institutional review board at the University of Washington (contact via Jane Edelson)) for researchers who meet the criteria for access to confidential data.

## Acknowledgements

The authors would also like to thank the Lighthouse Trust and its NCAP program team for their partnership in this research study.

## References

1. New report from UNAIDS shows that AIDS can be ended by 2030 and outlines the path to get there [Internet]. [cited 2023 Nov 3]. Available from: https://www.unaids.org/en/resources/presscentre/pressreleaseandstatementarchive/2023/july/unaids-global-aids-update

2. Long L, Kuchukhidze S, Pascoe S, Nichols BE, Fox MP, Cele R, et al. Retention in care and viral suppression in differentiated service delivery models for HIV treatment delivery in sub-Saharan Africa: a rapid systematic review. Journal of the International AIDS Society. 2020;23(11):e25640.

3. Kuo AP, Roche SD, Mugambi ML, Pintye J, Baeten JM, Bukusi E, et al. The effectiveness, feasibility and acceptability of HIV service delivery at private pharmacies in sub-Saharan Africa: a scoping review. Journal of the International AIDS Society. 2022;25(10):e26027.

4. Fatti G, Ngorima-Mabhena N, Mothibi E, Muzenda T, Choto R, Kasu T, et al. Outcomes of Three-Versus Six-Monthly Dispensing of Antiretroviral Treatment (ART) for Stable HIV Patients in Community ART Refill Groups: A Cluster-Randomized Trial in Zimbabwe. J Acquir Immune Defic Syndr. 2020 Mar 23;10.1097/QAI.0000000000002333.

5. Christ B, van Dijk JH, Nyandoro TY, Reichmuth ML, Kunzekwenyika C, Chammartin F, et al. Availability and experiences of differentiated antiretroviral therapy delivery at HIV care facilities in rural Zimbabwe: a mixed-method study. J Int AIDS Soc. 2022 Aug 25;25(8):e25944.

6. Sande O, Burtscher D, Kathumba D, Tweya H, Phiri S, Gugsa S. Patient and nurse perspectives of a nurse-led community-based model of HIV care delivery in Malawi: a qualitative study. BMC Public Health. 2020 May 14;20(1):685.

7. Bemelmans M, Baert S, Goemaere E, Wilkinson L, Vandendyck M, van Cutsem G, et al. Community-supported models of care for people on HIV treatment in sub-Saharan Africa. Tropical Medicine & International Health. 2014;19(8):968–77.

8. Pellecchia U, Baert S, Nundwe S, Bwanali A, Zamadenga B, Metcalf CA, et al. “We are part of a family”. Benefits and limitations of community ART groups (CAGs) in Thyolo, Malawi: a qualitative study. Journal of the International AIDS Society. 2017;20(1):21374.

9. Rasschaert F, Telfer B, Lessitala F, Decroo T, Remartinez D, Biot M, et al. A Qualitative Assessment of a Community Antiretroviral Therapy Group Model in Tete, Mozambique. PLOS ONE. 2014 Mar 20;9(3):e91544.

10. Tun W, Apicella L, Casalini C, Bikaru D, Mbita G, Jeremiah K, et al. Community-Based Antiretroviral Therapy (ART) Delivery for Female Sex Workers in Tanzania: 6-Month ART Initiation and Adherence. AIDS Behav. 2019 Sep 1;23(2):142–52.

11. Zakumumpa H, Rujumba J, Kwiringira J, Katureebe C, Spicer N. Understanding implementation barriers in the national scale-up of differentiated ART delivery in Uganda. BMC Health Services Research. 2020 Mar 17;20(1):222.

12. Adjetey V, Obiri-Yeboah D, Dornoo B. Differentiated service delivery: a qualitative study of people living with HIV and accessing care in a tertiary facility in Ghana. BMC Health Services Research. 2019 Feb 4;19(1):95.

13. Grimsrud A, Wilkinson L. Acceleration of differentiated service delivery for HIV treatment in sub-Saharan Africa during COVID-19. Journal of the International AIDS Society. 2021;24(6):e25704.

14. New report from UNAIDS shows that AIDS can be ended by 2030 and outlines the path to get there [Internet]. [cited 2023 Nov 3]. Available from: https://www.unaids.org/en/resources/presscentre/pressreleaseandstatementarchive/2023/july/unaids-global-aids-update

15. Feldacker C, Tweya H, Keiser O, Weigel R, Kalulu M, Fenner L, et al. Characteristics of adults and children diagnosed with tuberculosis in Lilongwe, Malawi: findings from an integrated HIV/TB clinic. Tropical Medicine & International Health. 2012;17(9):1108–16.

16. Huwa J, Tweya H, Mureithi M, Kiruthu-Kamamia C, Oni F, Chintedza J, et al. “It reminds me and motivates me”: Human-centered design and implementation of an interactive, SMS-based digital intervention to improve early retention on antiretroviral therapy: Usability and acceptability among new initiates in a high-volume, public clinic in Malawi. PLOS ONE. 2023 Jul 20;18(7):e0278806.

17. Makombe SD, Hochgesang M, Jahn A, Tweya H, Hedt B, Chuka S, et al. Assessing the quality of data aggregated by antiretroviral treatment clinics in Malawi. Bull World Health Organ. 2008 Apr;86(4):310–4.

18. Phiri S, Khan PY, Grant AD, Gareta D, Tweya H, Kalulu M, et al. Integrated tuberculosis and HIV care in a resource-limited setting: experience from the Martin Preuss centre, Malawi. Tropical Medicine & International Health. 2011;16(11):1397–403.

19. Phiri S, Neuhann F, Glaser N, Gass T, Chaweza T, Tweya H, et al. The path from a volunteer initiative to an established institution: evaluating 15 years of the development and contribution of the Lighthouse trust to the Malawian HIV response. BMC Health Services Research. 2017 Aug 9;17(1):548.

20. Tweya H, Ben-Smith A, Kalulu M, Jahn A, Ng’ambi W, Mkandawire E, et al. Timing of antiretroviral therapy and regimen for HIV-infected patients with tuberculosis: the effect of revised HIV guidelines in Malawi. BMC Public Health. 2014 Feb 20;14(1):183.

21. Tweya H, Feldacker C, Gadabu OJ, Ng’ambi W, Mumba SL, Phiri D, et al. Developing a point-of-care electronic medical record system for TB/HIV co-infected patients: experiences from Lighthouse Trust, Lilongwe, Malawi. BMC Res Notes. 2016 Mar 5;9:146.

22. Vorkas CK, Tweya H, Mzinganjira D, Dickie G, Weigel R, Phiri S, et al. Practices to improve identification of adult antiretroviral therapy failure at the Lighthouse Trust clinic in Lilongwe, Malawi. Tropical Medicine & International Health. 2012;17(2):169–76.

23. Orii L, Feldacker C, Tweya H, Anderson R. eHealth Data Security and Privacy: Perspectives from Diverse Stakeholders in Malawi. Proceedings of the ACM on Human-Computer Interaction. Forthcoming.

24. Braun V, Clarke V. Toward good practice in thematic analysis: Avoiding common problems and be(com)ing a knowing researcher. Int J Transgend Health. 24(1):1–6.

25. Elo S, Kääriäinen M, Kanste O, Pölkki T, Utriainen K, Kyngäs H. Qualitative Content Analysis: A Focus on Trustworthiness. SAGE Open. 2014 Jan 1;4(1):2158244014522633.

26. Okere NE, Meta J, Maokola W, Martelli G, Praag E van, Naniche D, et al. Quality of care in a differentiated HIV service delivery intervention in Tanzania: A mixed-methods study. PLOS ONE. 2022 Mar 15;17(3):e0265307.

27. Berner-Rodoreda A, Geldsetzer P, Bärnighausen K, Hettema A, Bärnighausen T, Matse S, et al. “It’s hard for us men to go to the clinic. We naturally have a fear of hospitals.” Men’s risk perceptions, experiences and program preferences for PrEP: A mixed methods study in Eswatini. PLOS ONE. 2020 Sep 23;15(9):e0237427.

28. Berner-Rodoreda A, Ngwira E, Alhassan Y, Chione B, Dambe R, Bärnighausen T, Phiri S, Taegtmeyer M, Neuhann F. “Deadly”,“fierce”,“shameful”: notions of antiretroviral therapy, stigma and masculinities intersecting men’s life-course in Blantyre, Malawi. BMC Public Health. 2021 Dec;21:1–3.

29. Mudavanhu M, West NS, Schwartz SR, Mutunga L, Keyser V, Bassett J, et al. Perceptions of Community and Clinic-Based Adherence Clubs for Patients Stable on Antiretroviral Treatment: A Mixed Methods Study. AIDS Behav. 2020 Apr 1;24(4):1197–206.

30. Mukumbang FC, Ndlovu S, van Wyk B. Comparing Patients’ Experiences in Three Differentiated Service Delivery Models for HIV Treatment in South Africa. Qual Health Res. 2022 Jan;32(2):238–54.

31. Walusaga HAG, Atuyambe LM, Muddu M, Mpirirwe R, Nangendo J, Kalibbala D, et al. Perceptions and factors associated with the uptake of the community client-led antiretroviral therapy delivery model (CCLAD) at a large urban clinic in Uganda: a mixed methods study. BMC Health Services Research. 2023 Oct 26;23(1):1165.

32. Ehrenkranz P, Grimsrud A, Holmes CB, Preko P, Rabkin M. Expanding the Vision for Differentiated Service Delivery: A Call for More Inclusive and Truly Patient-Centered Care for People Living With HIV. J Acquir Immune Defic Syndr. 2021 Feb 1;86(2):147–52.

33. Decroo T, Van Damme W, Kegels G, Remartinez D, Rasschaert F. Are Expert Patients an Untapped Resource for ART Provision in Sub-Saharan Africa? AIDS Research and Treatment. 2012 Apr 19;2012:e749718.

34. Sharer M, Davis N, Makina N, Duffy M, Eagan S. Differentiated Antiretroviral Therapy Delivery: Implementation Barriers and Enablers in South Africa. J Assoc Nurses AIDS Care. 2019;30(5):511–20.

